# A Retrospective Multi-Source Clinical Validation of Lenek Intelligent Radiology Assistant: An Artificial Intelligence-Based Chest Radiograph Screening and Triage System for High-Burden Pulmonary and Cardiac Conditions in India

**DOI:** 10.64898/2026.03.14.26348373

**Authors:** Vinit Singh, Ananya Jhamb, Shiladitya Sil, Sidesh Kumar, Chirag Agrawal, Anant Pareek, Aditya Gautam, Gurunath Parale, Shuchi Singh, Deepak Padmanabhan

**Affiliations:** Jawaharlal Nehru Medical College, KLE Academy of Higher Education & Research, Belagavi, Karnataka, India; Department of Oral Medicine and Radiology, North Bengal Dental College & Hospital, Thiknikata, West Bengal, India; Lenek Technologies Private Limited, Pune, Maharashtra, India; Department of Respiratory Medicine, Uttar Pradesh University of Medical Sciences, Saifai, Uttar Pradesh, India; Department of Cardiology, Ashwini Hospital, Solapur, Maharashtra, India; Department of Radiodiagnosis, Apollomedics Super Speciality Hospital, Lucknow, Uttar Pradesh, India; Department of Electrophysiology, Narayana Institute of Cardiac Sciences, Bengaluru, Karnataka, India

**Author notes:** **Corresponding author:** Vinit Singh, Jawaharlal Nehru Medical College, KLE Academy of Higher Education and Research, Belagavi, Karnataka (590010).

**Keywords:** Artificial Intelligence in Radiology, Chest-radiography, Tuberculosis screening, AI-driven triage, Access to healthcare, Screening tool

## Abstract

**Background:** A critical radiologist shortage exists in India, leading to delayed chest radiograph (CXR) interpretation. This leads to disease progression, higher morbidity, and mortality. Artificial intelligence-based CXR interpretation by Lenek Intelligent Radiology Assistant (LIRA) is a promising solution. This study aims to establish the screening and triaging capabilities of LIRA by assessing its accuracy in detecting abnormalities and pathologies in CXRs from geographically diverse institutions.

**Methods:** We conducted a retrospective multi-source validation of the diagnostic accuracy of LIRA for the detection of general abnormalities, tuberculosis, consolidation, pleural effusion, pneumothorax, and cardiomegaly. De-identified chest radiographs were input into LIRA models. The obtained interpretations were compared to the established ground truth reporting for the calculation of sensitivity, specificity, and AUROC with 95% CI for individual pathologies across varying probability thresholds.

**Results:** LIRA demonstrated high sensitivity for general abnormality detection (AUROC 0.93-0.986, 84.4-97.1% sensitivity, 88.9-92.4% specificity) and tuberculosis triaging (Shenzhen & Montgomery: 88.5-89.7% sensitivity, 89.9-90.5% specificity; Jaypee: 98.7% sensitivity, 63.6% specificity). For consolidation (AUROC 0.884-0.895, 96.4-96.9% sensitivity, 70.8-77.1% specificity), pleural effusion (AUROC 0.942-0.967, 79.7-99.1% sensitivity, 81.2-87.7% specificity), pneumothorax (AUROC 0.87, 90.6-94.8% sensitivity, 79.5-82.7% specificity) and cardiomegaly (AUROC 0.883, 95.1% sensitivity, 81.6% specificity), the model exhibited commendable accuracy as well.

**Conclusions:** The diagnostic performance of LIRA was consistent across various pathologies and chest radiographs from diverse geographic locations, with particular strengths in abnormality detection and tuberculosis screening. The risk-stratified triaging and high sensitivity of LIRA make it a reliable adjunct solution to address radiologist shortages, reduce turnaround times, and support India’s tuberculosis elimination goals.

## INTRODUCTION

Chest radiography is the most widely performed imaging investigation for cardiopulmonary diseases worldwide. Chest X-Rays (CXRs) serve as the first-line diagnostic modality for tuberculosis, pneumonia, pleural diseases, pneumothorax, and cardiomegaly. Driven by the country’s substantial burden of tuberculosis, chronic respiratory diseases, and cardiovascular conditions, India performs millions of chest radiographs annually. However, due to radiologist shortages, this burden of CXRs does not meet the timely and quality interpretation.^1^ The Indian healthcare infrastructure today caters to a population exceeding 1.4 billion. India experiences a severe diagnostic bottleneck with an estimated radiologist-to-population ratio of 1:100,000, which adversely affects patient outcomes.^2^ This problem further exaggerates in tier-2, tier-3, and rural healthcare settings, where there is a larger shortage of radiologists, and patients often travel long distances to reach a tertiary care center, resulting in delayed diagnostics.^3^

India contributes nearly one-quarter of the global tuberculosis burden with approximately 2.6 million cases annually and has set an ambitious target to eliminate tuberculosis by 2030, five years ahead of the global Sustainable Development Goal timeline.^4, 5^ Delayed radiological interpretation of conditions like tuberculosis, consolidation, and pneumothorax leads to disease progression, increased morbidity, higher mortality rates, and continued disease transmission in communities.^6, 7^

Deep learning-based image analysis by Artificial Intelligence (AI) is a promising solution to address these challenges. These AI algorithms can rapidly analyze chest radiographs and flag abnormal studies, therefore assisting clinicians in prioritizing cases that require urgent attention.^8, 9^ AI tools have demonstrated high diagnostic accuracy; however, many remain limited to single-disease detection or lack validation across heterogeneous datasets that are representative of real-world clinical practice.^10, 11^

LIRA (Lenek Intelligent Radiology Assistant) (Lenek Technologies Private Limited, Pune, India) is an AI-enabled radiological computer-assisted triage, interpretation, and reporting software application. It is designed to assist qualified healthcare professionals in the review and analysis of adult CXR scans (AP/PA views) for the presence of abnormalities, multiple high-impact thoracic and cardiac pathologies, including tuberculosis, pulmonary consolidation, pleural effusion, pneumothorax, and cardiomegaly. LIRA also has a risk-stratified output for tuberculosis suspicion, where the predictions are scored by a quantitative “Tuberculosis Score” (0-100%) and categorized by a color-coded scoring system into red (high likelihood), yellow (indeterminate), and green (low likelihood) zones, thus enabling rational triage and referral decisions in resource-limited settings.

This is a multi-source retrospective study aimed at the clinical validation of the screening and triaging capabilities of LIRA across diverse publicly available CXR datasets from international institutions (National Institutes of Health (NIH); Department of Health and Human Services, Montgomery County; Shenzhen No.3 People’s Hospital, Guangdong Medical College, Shenzhen; CheXpert, Stanford Hospital; Society for Imaging Informatics in Medicine, American College of Radiology (SIIM-ACR)) and Indian institutions (Jaypee University of Information Technology) to assess the diagnostic accuracy, generalizability, and clinical feasibility of LIRA-CXR interpretation and reporting as a solution to bridge the radiology reporting gap and support tuberculosis elimination efforts in India and other high-burden, low-resource environments.

## METHODS

### Study Design and Ethical Considerations

This was a multi-source retrospective clinical validation study aimed to evaluate the diagnostic accuracy of LIRA in screening and triaging of CXRs for general abnormalities, tuberculosis, consolidation, pleural effusion, pneumothorax, and cardiomegaly. The study protocol was designed in adherence to the Standards for Reporting of Diagnostic Accuracy Studies (STARD) 2015 guidelines. Ethical approval was not required for this study as it involved the retrospective analysis of de-identified, publicly available CXR datasets (NIH, CheXpert, Montgomery, Shenzhen, SIIM-ACR, and Jaypee University of Information Technology). No human subjects were directly recruited, and no identifiable private information was accessed. The study was conducted in compliance with the Health Insurance Portability and Accountability Act (HIPAA) and the Indian Council of Medical Research (ICMR) National Ethical Guidelines for Biomedical and Health Research.

### Study Protocol

The study used de-identified CXRs obtained from geographically diverse publicly available CXRs from international institutions, which were input into the LIRA Models for interpretation. The obtained LIRA interpretation of the CXRs was then compared to the established ground truth. The ground truth (reference standard) was established by consensus labelling of CXRs by board-certified radiologists. Based on this comparison, the performance metrics were calculated to establish the diagnostic accuracy of LIRA.

### Eligibility Criteria

This study included frontal CXR’s (Posteroanterior [PA] or Anteroposterior [AP] views) from adult patients (>18 years). Images of the pediatric population, non-CXR modalities (CT, MRI), and images with significant artifacts or metallic implants obscuring the lung fields were excluded from the study.

### Data Acquisition

The clinical validation was performed on five distinct CXR datasets from global benchmark institutes to establish the diagnostic accuracy of the LIRA across geographically diverse patient populations and imaging equipment.

1. **National Institutes of Health (NIH, USA):** We used a subset of the NIH ChestX-ray8 dataset ^12^ for general abnormality detection, comprising 1,344 chest radiographs (653 abnormal, 691 normal) with ground truth established by consensus of three American Board of Radiology-certified radiologists. Another subset of 810 chest radiographs labeled by five board-certified radiologists was used for specific pathologies (83 consolidation, 226 pleural effusion, 136 pneumothorax, 81 cardiomegaly).
2. **CheXpert (USA):** It is a large public dataset consisting of 224,316 chest radiographs of 65,240 patients collected from Stanford Hospital between 2002 and 2017.^13^ A subset consisting of 234 studies was labelled by radiologists, among which 202 of these images are of AP/PA view, which were used for testing here (32 consolidation, 64 pleural effusion).
3. **Montgomery County (USA) & Shenzhen (China):** These datasets were created by the National Library of Medicine in collaboration with the Department of Health and Human Services, Montgomery County, Maryland, USA & Shenzhen No. 3 People’s Hospital, Guangdong Medical College, Shenzhen, China.^14^ These served as the primary benchmarks for Tuberculosis (TB) detection in our study. The Shenzhen dataset consisted of 662 images: 269 of confirmed TB positive cases, and 326 of confirmed non-TB. The Montgomery dataset consisted of 137 images: 58 of confirmed TB positive, and 79 of confirmed TB negative.
4. **Society for Imaging Informatics in Medicine - American College of Radiology (SIIM-ACR)** A large-scale open-source dataset comprising 3,286 pneumothorax-positive and 8,296 pneumothorax-negative images was utilized for pneumothorax validation.^15^
5. **Jaypee University of Information Technology (India):** A publicly available dataset from Jaypee University of Information Technology ^16^ with 154 images (77 confirmed TB positive, 77 confirmed TB negative) was used to assess the TB model’s sensitivity in the Indian demographic.

### Lenek Intelligent Radiology Assistant (LIRA) Overview

LIRA (Lenek Intelligent Radiology Assistant) v1.0 is an AI-enabled radiological computer-assisted triage, interpretation, and reporting software as a Medical Device (SaMD). It is classified as a Class B Medical Device under India’s Medical Device Rules (MDR) 2017. It is designed to assist qualified healthcare professionals in the review and analysis of adult chest X-ray (CXR) scans (AP/PA views). LIRA is intended for use by trained Radiologists, Referring Physicians, and Radiographers as an assistive tool. It aids in the triage and prioritization of CXR studies by identifying and highlighting potentially critical findings, thereby reducing reporting turnaround times in teleradiology and hospital Picture Archiving and Communication System (PACS) environments, thereby streamlining radiological workflows. The software analyzes CXR studies to identify the presence of general abnormalities, calculate a Tuberculosis (TB) probability score (0-100%) for screening purposes, and detect specific critical findings, including Consolidation, Pneumothorax, Pleural Effusion, and Cardiomegaly. It also produces color heatmap overlays that highlight image regions potentially relevant to the identified findings.

#### Abnormality Detection Model

The Abnormality detection model reports under a binary classification of "Abnormal/ Yes" or "Normal/ No". "Abnormal/ Yes” indicates that the model has detected features that deviate from what it considers normal. This suggests that the study may warrant further review. "Normal/ No” means that the AI model did not detect features it considers abnormal.

#### Tuberculosis (TB) Triage Model

For every radiograph that LIRA screens, the model assesses specific ‘Tuberculosis tags’ (radiological indicators of active or prior infection) such as cavitation, infiltration, nodules, and pleural effusion to derive a quantitative TB probability score between 0-100% to assess the probability of a patient harboring TB (Figure 1).

**FIGURE 1:**
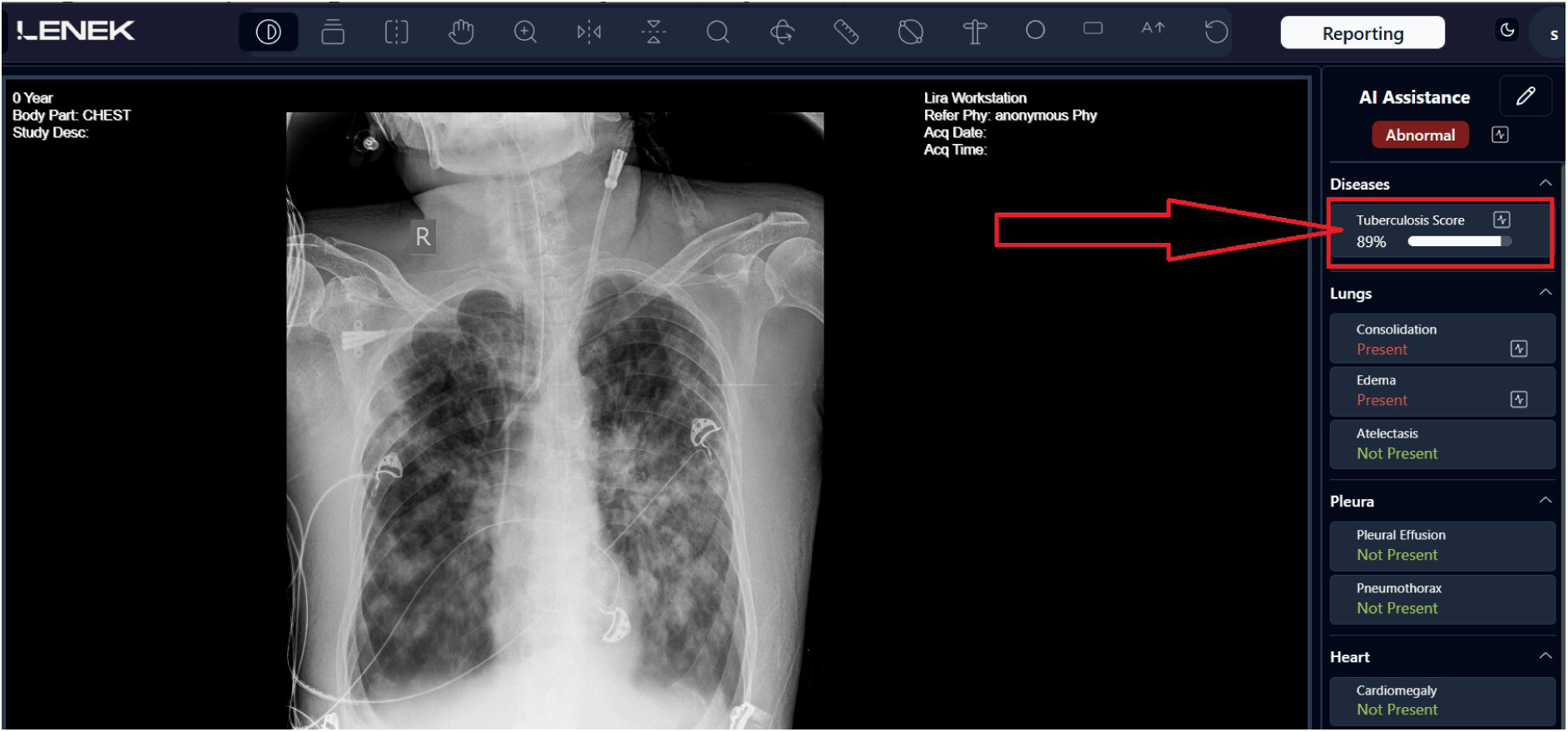
The TB Score Generated by LIRA for Abnormal CXRs with Probable TB. For every radiograph that LIRA screens, it assesses specific radiological indicators (cavitation, infiltration, nodules, pleural effusion) to derive a quantitative TB probability score (0-100%).

Based on the quantitative score, LIRA also gives a risk-stratified output for TB suspicion using a color-coded zone system, providing an immediate visual assessment of TB suspicion (Figure 2)

**FIGURE 2:**
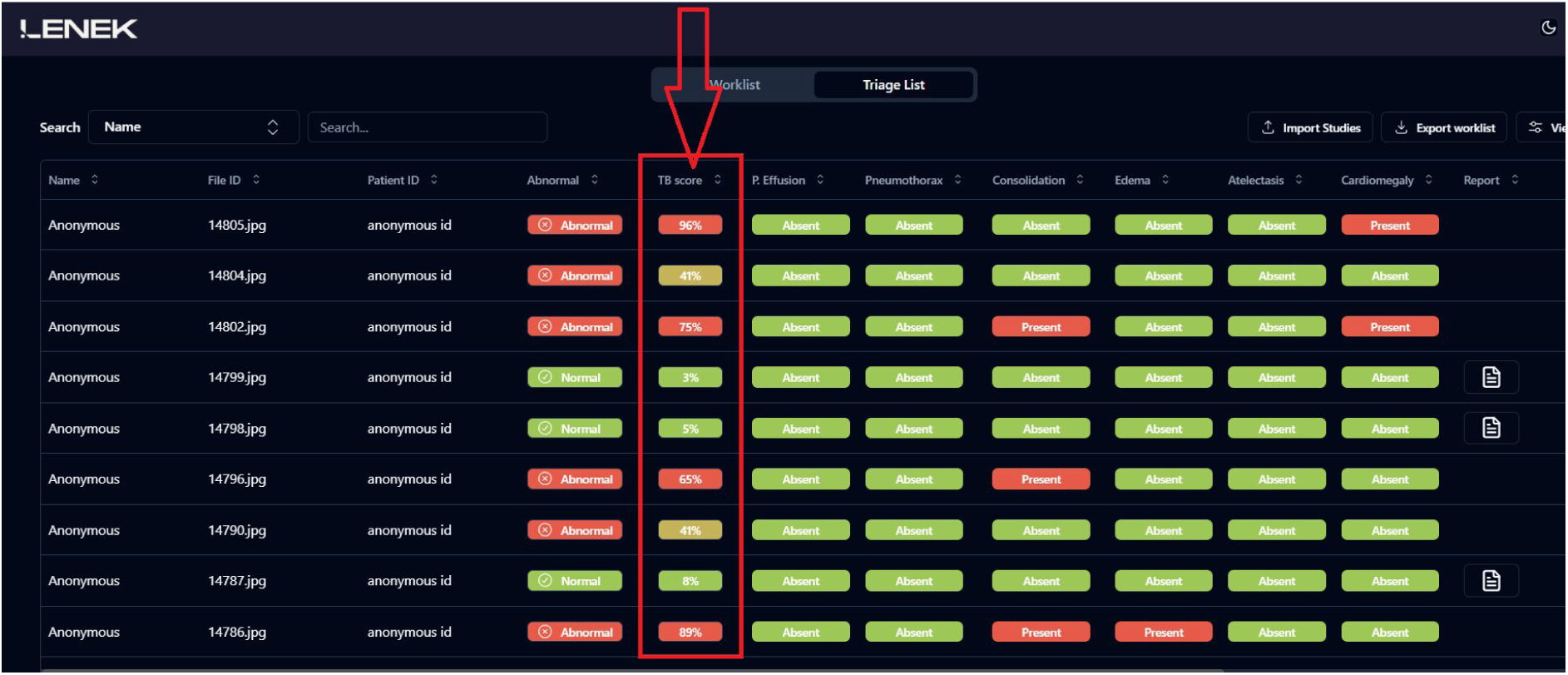
LIRA Interface and the Unique Three-Zone Triage Framework for TB Screening. The screenshot demonstrates the LIRA user interface displaying the Triage List with color-coded risk stratification (Green, Yellow, Red) for tuberculosis suspicion alongside binary outputs for other pathologies.

#### Interpretation of Color Coding by LIRA

The Green Zone (Score 0-X%) indicates a low level of suspicion for TB. This means that, according to the AI analysis, cases in this category are unlikely to be indicative of Tuberculosis. The Red Zone (Score Y-100%) signifies a high level of suspicion for TB. This means that the AI model has identified features consistent with the disease. The Yellow Zone (Score X+1 to Y-1%) represents an intermediate level of suspicion. Interpretation requires further consideration in conjunction with the output from the "Abnormality Detection Model", which is a crucial determinant in refining the suspicion level for Tuberculosis when the TB model yields a Yellow Zone (Figure 3)

**FIGURE 3:**
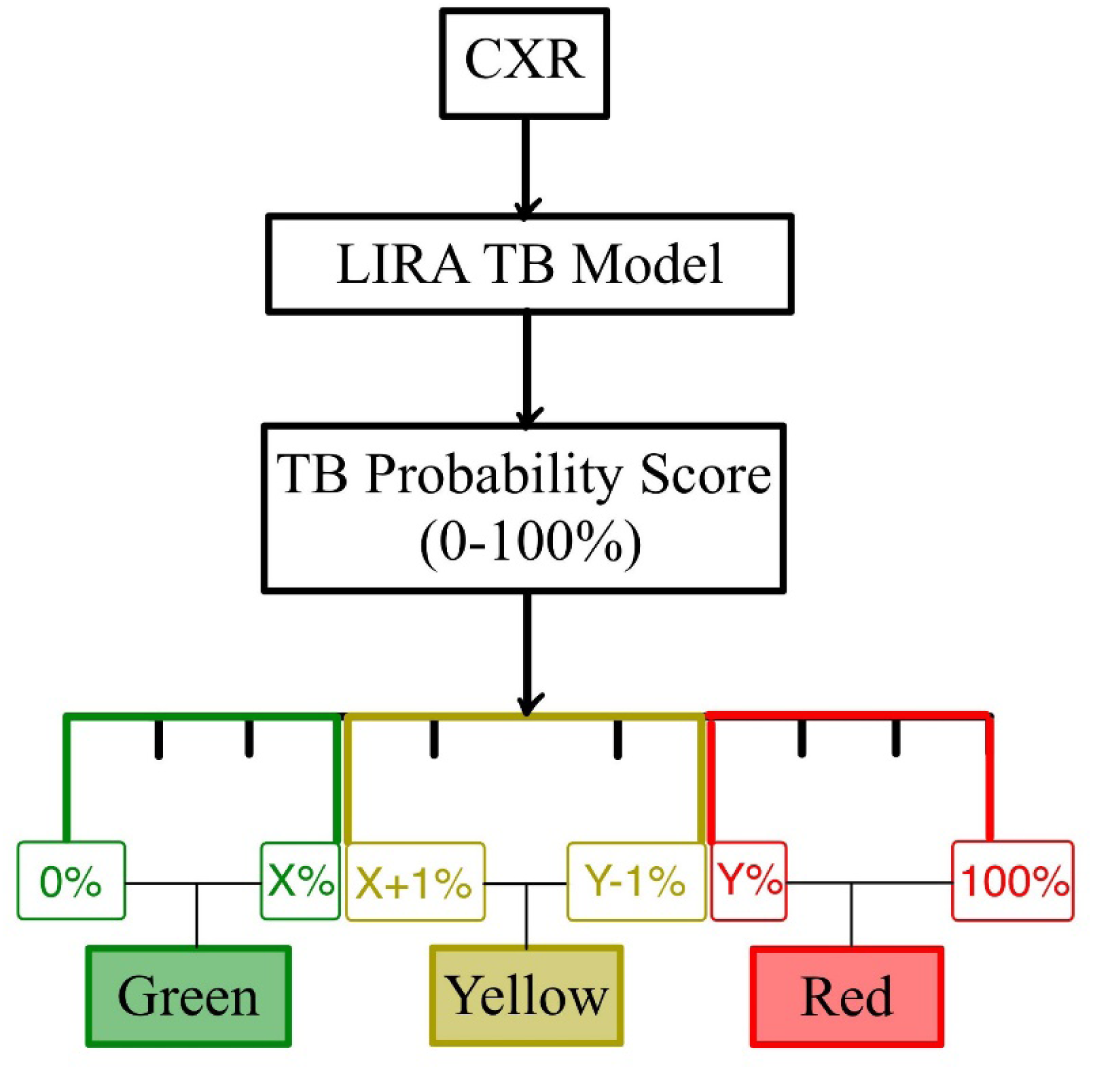
Color Coding by LIRA for Tuberculosis Triaging. The diagram illustrates the risk-stratified output system. X% represents the upper limit of the Green Zone (low suspicion), and Y% represents the lower limit of the Red Zone (high suspicion).

#### Pathology Detection Model for other Pathologies

LIRA also includes models for detecting specific pathologies such as Consolidation, Pleural Effusion, Pneumothorax, and Cardiomegaly. The system uses a binary classification model ("Yes/Present" or "No/Not Present") accompanied by heatmap overlays to help radiologists in localizing the pathology. A heatmap is an overlay on the medical image that visually highlights the specific regions the AI model identified as most indicative of the detected pathology (Figure 4). These models are designed as assistive tools to provide supplementary information to radiologists.

**FIGURE 4:**
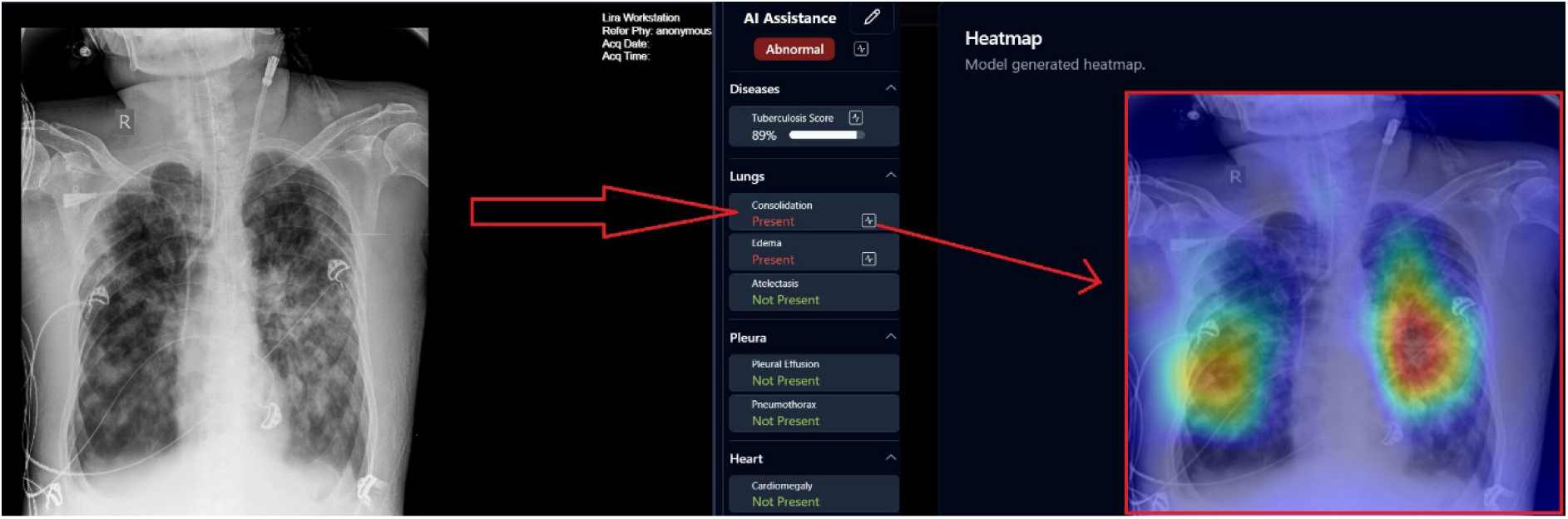
A Positive Case of Lung Consolidation in a Chest X-ray (PA View) The left image depicts the actual radiograph, and the right image depicts the heatmap generated by LIRA to highlight the area of interest (consolidation) to assist radiologists in localization.

### Reference Standard and Ground Truth establishment

The reference standard was strictly defined for each dataset to ensure reliability. For the NIH dataset, labels were determined by the consensus of three U.S. board-certified radiologists. For the CheXpert validation set, labels were provided by board-certified radiologists using the Fleischner Society definitions. For the Shenzhen, Montgomery, and Jaypee datasets, labels were based on microbiological confirmation and expert clinical diagnosis as provided in the source dataset documentation.

### Statistical Analysis and Performance Metrics

Diagnostic accuracy of LIRA was assessed by calculating Sensitivity, Specificity, and the Area Under the Receiver Operating Characteristic (AUROC), with 95% confidence intervals (CI) calculated using De-Long’s method for AUROC and Wilson score method for sensitivity and specificity. For the TB triage model, sensitivity was defined as the proportion of true positive cases classified correctly into "Red" or "Yellow" categories. Specificity was defined as the proportion of true negatives classified correctly into the "Green" category. These metrics were calculated across multiple specified probability thresholds (e.g., 0.4, 0.5, 0.6) to identify the optimal operating point for a triage setting with high sensitivity to minimize missed diagnoses.

## RESULTS

### Results of Abnormality Detection Model

The abnormality detection model achieved excellent diagnostic performance across the NIH and the Montgomery datasets. On the NIH dataset (1,344 images), AUROC was 0.986 (95% CI: 0.98-0.992). At the threshold of 0.5, the model exhibited 97.1% sensitivity (CI: 96.2-98.0%) and 88.9% specificity (CI: 87.2- 90.5%). Higher thresholds (0.6) improved specificity to 93.6% while maintaining sensitivity above 95%. In the Montgomery dataset (137 images), the calculated AUROC was 0.93 (CI: 0.88-0.971). At threshold 0.5, sensitivity was 84.4% (CI: 78.4-90.5%) with a specificity of 92.4% (CI: 86.5-96.0%) (Figure 5).

**FIGURE 5:**
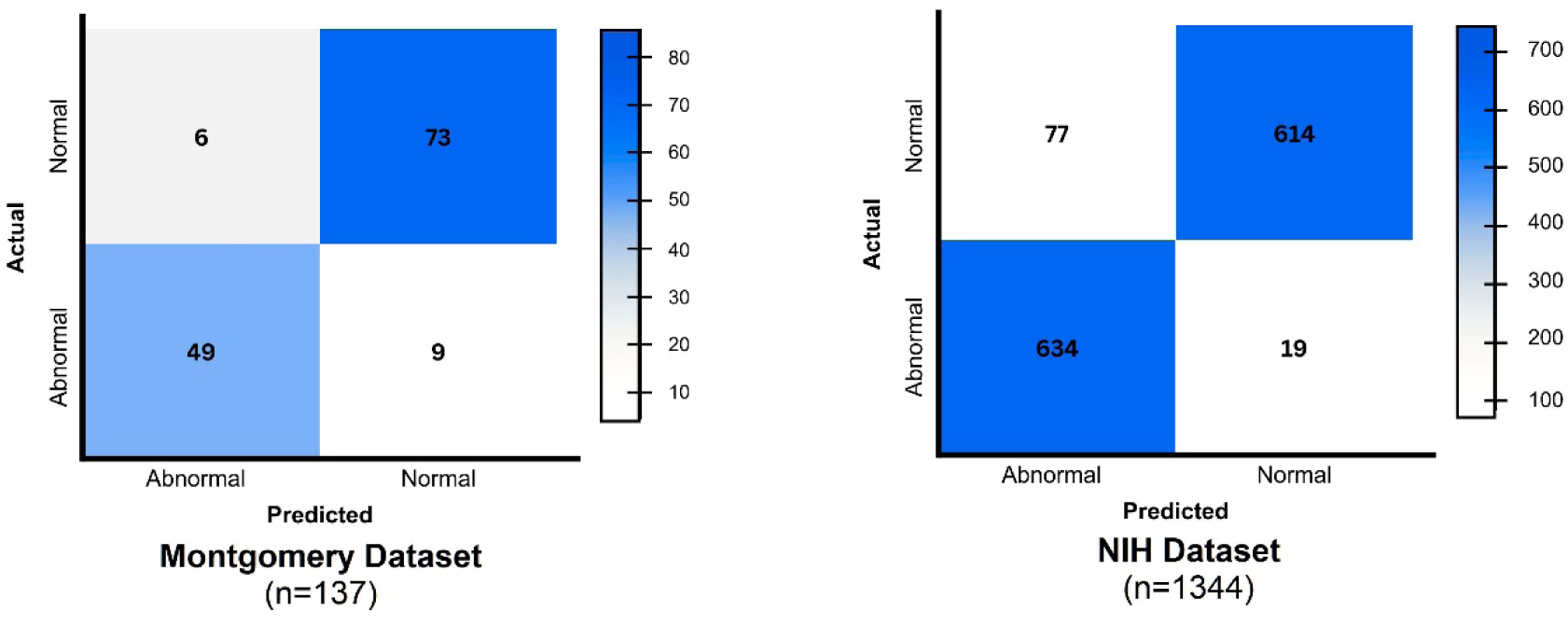
Confusion Matrix for Abnormality Detection Model. The figure illustrates the diagnostic performance of the LIRA abnormality detection model across two datasets. The left panel displays the confusion matrix for the Montgomery dataset (n=137), and the right panel displays the confusion matrix for the NIH dataset (n=1,344). The axes represent Actual (y-axis) versus Predicted (x-axis) classifications for ‘Abnormal’ and ‘Normal’ cases.

### Results of Tuberculosis Triage Model

The TB model demonstrated consistently high sensitivity across three geographically diverse datasets using the three-zone triage classification system. In the Shenzhen Dataset (595 images), LIRA correctly identified 238 of 269 TB-positive cases (red: 216, yellow: 22) and achieved a sensitivity of 88.5%. Specificity reached 90.5% with 295 of 326 TB-negative cases appropriately classified into the “Green” category. In the Montgomery Dataset (137 images), the model correctly classified 52 of 58 TB-positive cases (red: 50, yellow: 2) with a sensitivity of 89.65%. Calculated specificity was 89.9% (correctly identified 71 of 79 TB-negative cases as green), demonstrating consistent performance despite a smaller sample size. The model demonstrated the highest sensitivity (98.7%, correctly identified 76 of 77 TB-positive cases) in the Jaypee Dataset (154 images), though specificity was 63.6% (49 of 77 TB-negative cases classified “green”), possibly reflecting different disease severity or imaging characteristics in this Indian population (Figure 6).

**FIGURE 6:**
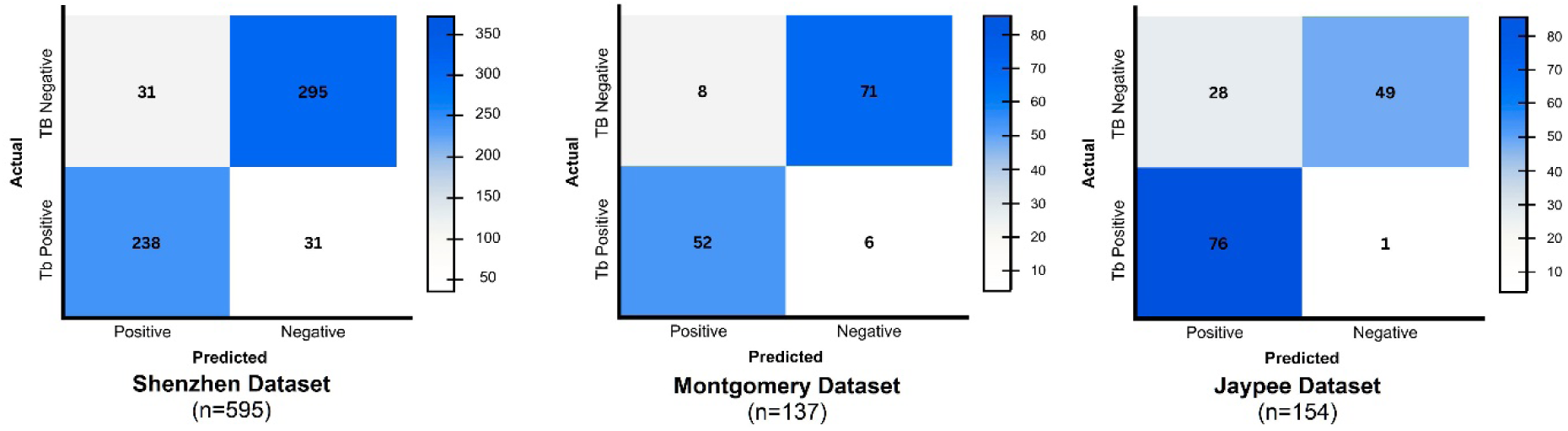
Confusion Matrices for Tuberculosis Triage Model. The figure illustrates the performance of the TB Triage Model across three datasets: Shenzhen (left), Montgomery (center), and Jaypee (right).

### Results of Consolidation Detection Model

In the CheXpert Dataset (202 images), the calculated AUROC was 0.894 (CI: 0.85-0.936). Sensitivity reached 96.9% (CI: 94.5-99.3%) with a specificity of 77.1% (CI: 71.2-82.8%) at the threshold of 0.55. In the NIH Dataset (810 images), the model achieved an AUROC of 0.885 (CI: 0.863-0.907), with a sensitivity of 96.4% (CI: 95.1-97.7%) and a specificity of 70.8% (CI: 67.7-74.0%) at a threshold of 0.55 (Figure 7).

**FIGURE 7:**
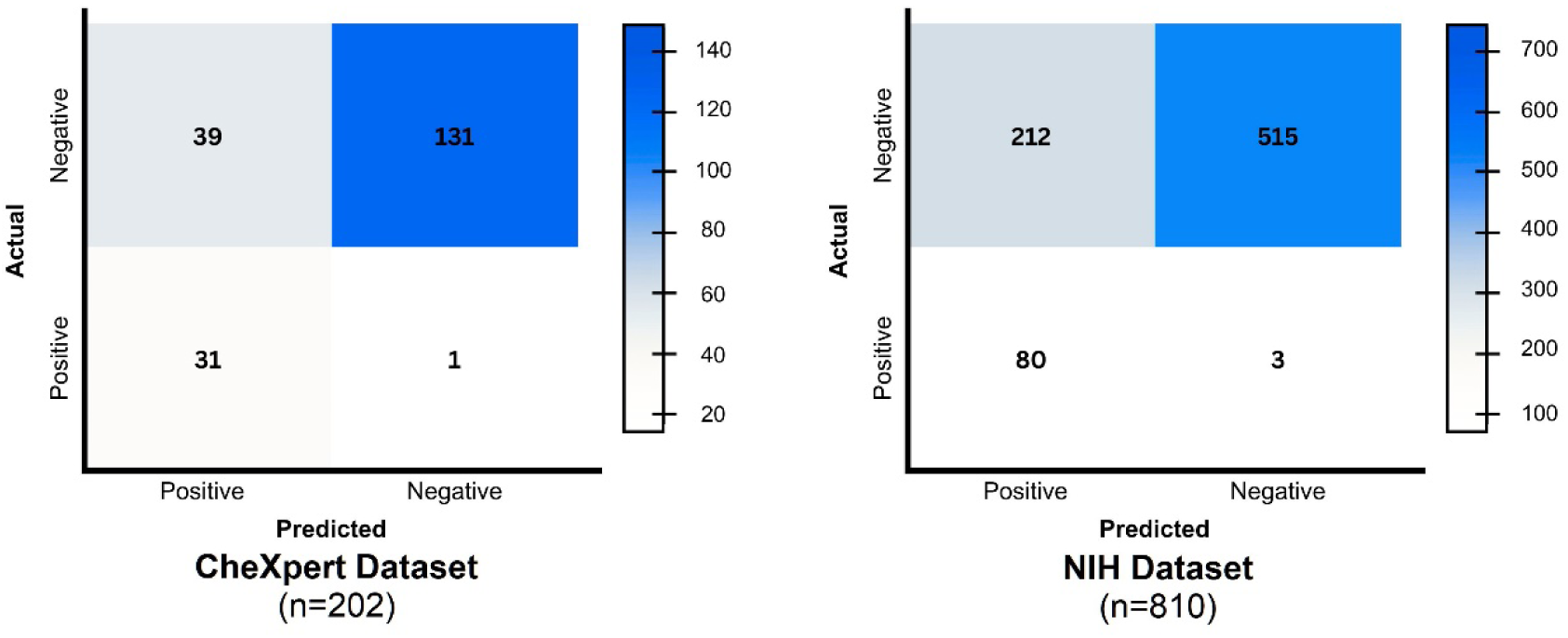
Confusion Matrices for Consolidation Detection Model. Performance of the consolidation detection model on the CheXpert dataset (left) and the NIH dataset (right).

### Results Of Pleural Effusion Detection Model

In the NIH Dataset (810 images), LIRA achieved a high AUROC of 0.967 (CI: 0.95-0.98), with a sensitivity of 99.1% (CI: 98.47-99.76%) and a specificity of 81.2% (CI: 78.5-83.8%) at a threshold of 0.45. In the CheXpert Dataset (202 images), AUROC was 0.942 (CI: 0.91-0.974), with a sensitivity of 79.7% (CI: 74.1-85.2%) and a specificity of 87.7% (CI: 83.1-92.2%) at a threshold of 0.45 (Figure 8).

**FIGURE 8:**
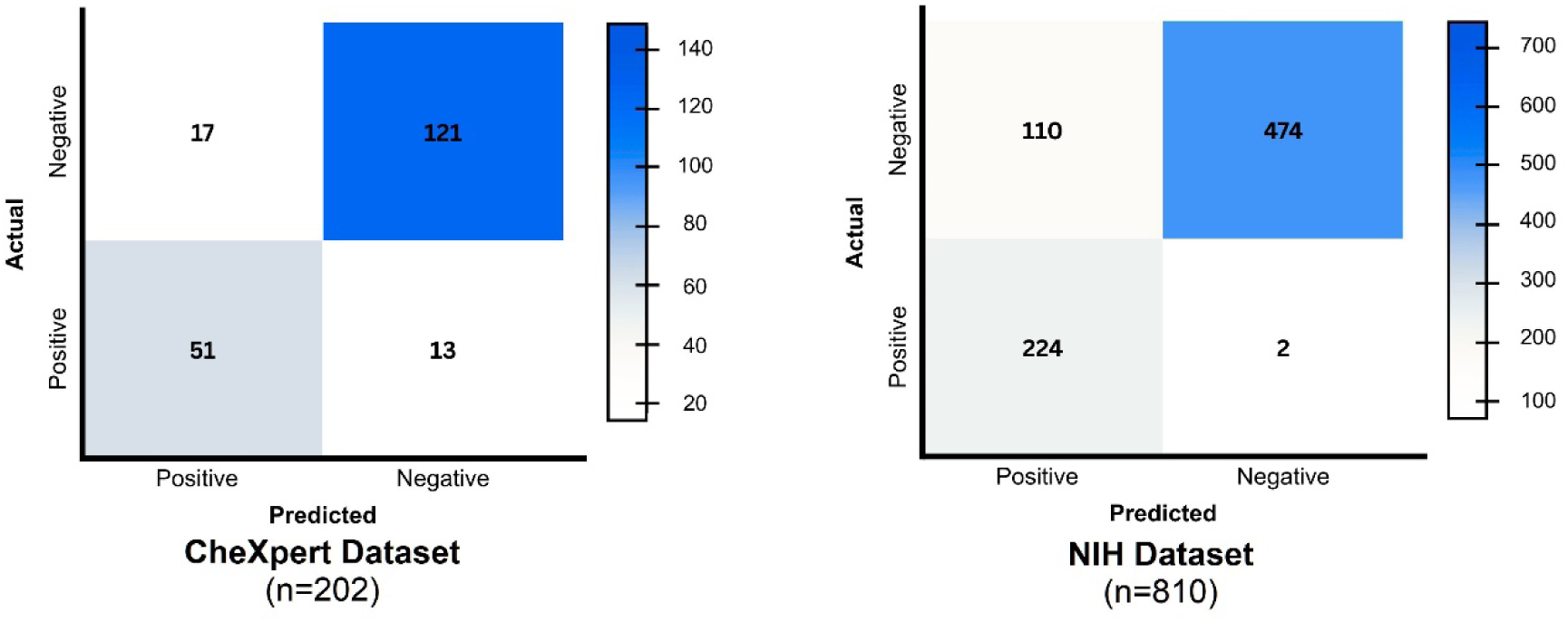
Confusion Matrices for Pleural Effusion Detection Model. Comparison of model performance for detecting pleural effusion on the CheXpert dataset (left) and the NIH dataset (right).

### Results of Pneumothorax Detection Model

In the NIH Dataset (810 images), an AUROC of 0.872 (CI: 0.849-0.895), sensitivity of 94.8% (CI: 93.3-96.4%), and specificity of 79.5% (CI: 76.7-82.3%) were achieved at a threshold of 0.38. In the SIIM-ACR Dataset (11,582 images), an AUROC of 0.867 (CI: 0.86-0.873), sensitivity of 90.6% (CI: 90.13-91.2%), and specificity of 82.7% (CI: 82.0-83.38%) were observed at a 0.38 threshold. The large sample size provided narrow confidence intervals, demonstrating stable performance of the model (Figure 9).

**FIGURE 9:**
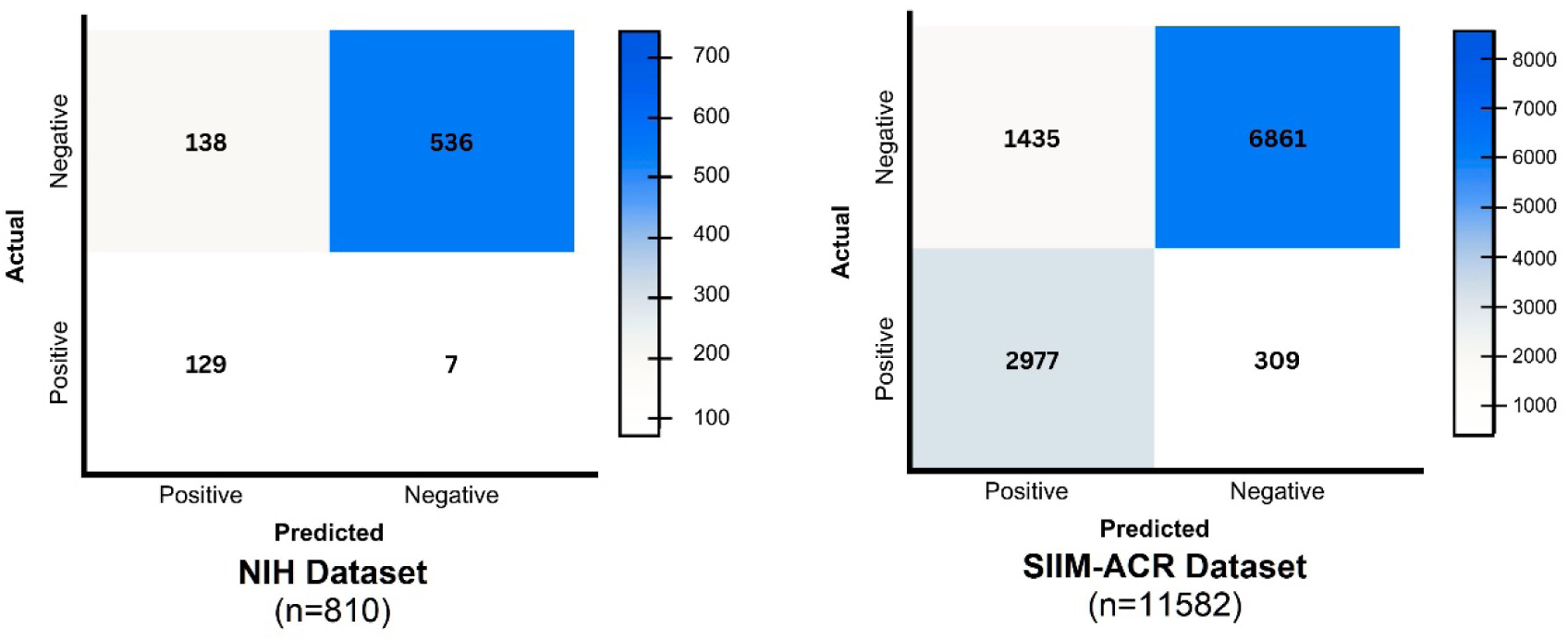
Confusion Matrices for Pneumothorax Detection Model. Performance metrics for pneumothorax detection on the NIH dataset (left) and the large-scale SIIM-ACR dataset (right).

### Results of Cardiomegaly Detection Model

In the NIH Dataset (810 images), AUROC = 0.883 (CI: 0.861-0.905), sensitivity of 95.1% (CI: 93.5-96.55%), and specificity of 81.6% (CI: 78.95-84.28%) were achieved at a threshold of 0.39 (Figure 10).

**FIGURE 10:**
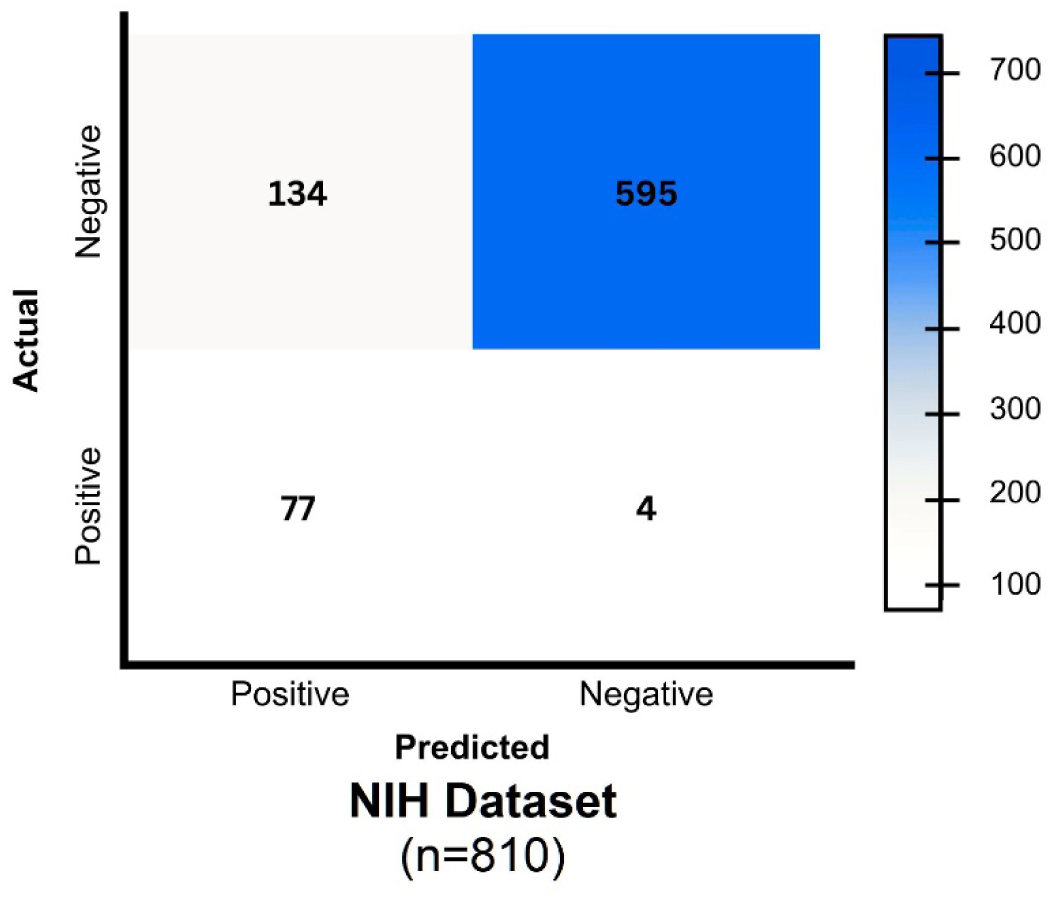
Confusion Matrix for Cardiomegaly Detection Model. The matrix displays the classification performance of LIRA for cardiomegaly detection on the NIH dataset.

### Summary of Findings

A comprehensive summary of the diagnostic performance of LIRA across all studied pathologies and datasets is presented below. These metrics highlight the model’s consistent sensitivity and specificity across diverse geographic populations and imaging equipment, supporting its utility in high-burden settings (Figure 11, Table 1).

**FIGURE 11:**
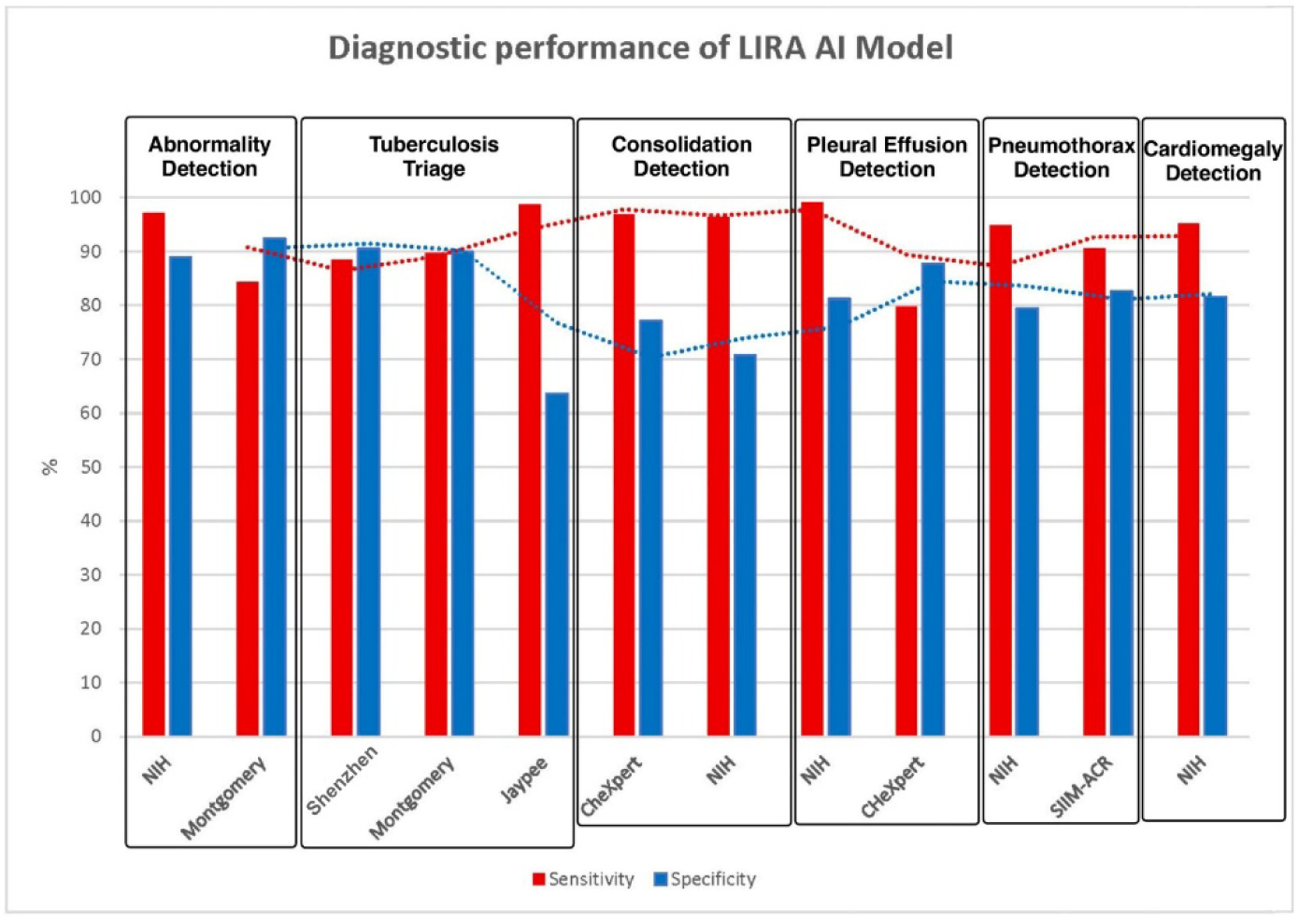
Sensitivity and Specificity of LIRA AI Model Across Chest X-ray Datasets. A comparative bar chart summarizing the sensitivity (red) and specificity (blue) of LIRA across all tested pathologies (Abnormality, Tuberculosis, Consolidation, Pleural Effusion, Pneumothorax, Cardiomegaly) and datasets.

**Table 1.** Diagnostic Performance of Lira AI Model Across Chest X-Ray Datasets. The table summarizes the diagnostic accuracy (Sensitivity, Specificity, AUROC) of the LIRA model for detecting general abnormalities, tuberculosis, consolidation, pleural effusion, pneumothorax, and cardiomegaly across heterogeneous datasets. Abbreviations: AUROC: Area Under the Receiver Operating Characteristic; CI: Confidence Interval; NIH: National Institutes of Health; SIIM-ACR: Society for Imaging Informatics in Medicine - American College of Radiology; ED: Emergency Department; TB: Tuberculosis; NA: Not Applicable.

| **Sr No** | **Parameter** | **Dataset** | **Sample size (positive/negative)** | **AUROC(95% CI)** | **Optimum threshold** | **Sensitivity (%)** | **Specificity (%)** | **Intended clinical role** |
| --- | --- | --- | --- | --- | --- | --- | --- | --- |
| 1. | Abnormality detection | NIH (USA) | 1344 (653/691) | 0.986 (0.98-0.992) | 0.5 | 97.1 | 88.9 | Rapid screening & workflow reduction |
| 2. | Abnormality detection | Montgomery (USA) | 137 (58/79) | 0.93 (0.88-0.971) | 0.5 | 84.4 | 92.4 | Primary triage |
| 3. | Tuberculosis | Shenzhen (China) | 595 (269/326) | NA | 0.2, 0.6 | 88.5 | 96.9 | TB Screening |
| 4. | Tuberculosis | Montgomery (USA) | 137 (58/79) | NA | 0.2, 0.6 | 89.65 | 98.7 | TB Screening |
| 5. | Tuberculosis | Jaypee (India) | 154 (77/77) | NA | 0.2, 0.6 | 98.7 | 85.7 | TB screening |
| 6. | Consolidation | CheXpert (USA) | 202 (32/170) | 0.894 (0.85-0.936) | 0.55 | 96.9 | 77.1 | Pneumonia triage |
| 7. | Consolidation | NIH (USA) | 810 (83/727) | 0.885 (0.86-0.91) | 0.55 | 96.4 | 70.8 | ED prioritization |
| 8. | Pleural effusion | NIH (USA) | 810 (226/584) | 0.967 (0.95-0.98) | 0.45 | 99.1 | 81.2 | Urgent case flagging |
| 9. | Pleural effusion | CheXpert (USA) | 202 (64/138) | 0.942 (0.91-0.974) | 0.45 | 79.7 | 87.7 | In-patient triage |
| 10. | Pneumothorax | NIH (USA) | 810 (136/674) | 0.872 (0.849-0.895) | 0.38 | 94.8 | 79.5 | Emergency detection |
| 11. | Pneumothorax | SIIM-ACR | 11582 (3286/8296) | 0.867 (0.86-0.873) | 0.38 | 90.6 | 82.7 | Emergency triage |
| 12. | Cardiomegal<br>y | NIH (USA) | 810 (81/<br>729) | 0.883<br>(0.861-0.905) | 0.39 | 95.1 | 81.6 | Chronic<br>disease flag |

## DISCUSSION

This multi-source clinical validation study shows that LIRA achieves comparable diagnostic performance across heterogeneous datasets and imaging environments. This supports its intended role as a clinical screening and triage tool in resource-limited healthcare settings. These findings have important implications for catering to systemic gaps in India’s healthcare delivery systems and for supporting national tuberculosis (TB) elimination efforts.

### Addressing Healthcare Workforce Shortages and Diagnostic Delays

India faces a critical shortage of radiologists, resulting in substantial diagnostic delays. Such delays adversely affect patient outcomes. With approximately one radiologist per 100,000 population, compared to 10 to 15 per 100,000 in high-income countries, the imbalance between imaging acquisition and interpretation capacity continues to widen.^1, 2^ This deficit translates into prolonged turnaround times in terms of reporting, often ranging from several days in district hospitals to weeks in primary health centers, during which treatable conditions may worsen, and infectious diseases may continue to spread.^3^ By cutting reporting delays from days to minutes, LIRA’s capacity to provide near-real-time preliminary interpretations radically alters this paradigm. When deployed as a first-line screening tool, LIRA can prioritize studies with critical findings for immediate radiologist review while also clearing likely to be normal examinations. This focused allocation of scarce specialized resources allows radiologists to completely focus on complex cases, accelerate triage decisions in emergencies, and minimize delays in outpatient follow-up.^8, 17^ The economic impact of such efficiency gains is substantial. Diagnostic delays lead to higher healthcare costs, prolonged hospital stays, advanced disease management, loss of productivity and livelihood, and increased transmission of communicable diseases. By facilitating earlier diagnosis and timely intervention, AI-assisted screening redirects healthcare expenditure toward prevention and early treatment. Such approaches are more cost-effective than managing advanced disease. ^18, 19^

### Tuberculosis Screening, Triage, and Elimination Strategy

Tuberculosis continues to remain one of India’s most challenging public health diseases. Despite ongoing efforts under the National Tuberculosis Elimination Program (NTEP), India continues to contribute approximately 27-28% of global TB cases, with an estimated incidence of 2.6 million and a mortality of nearly 450,000 deaths annually.^4, 5^ Delayed diagnosis, under-reporting, and failure to follow-up remain the primary factors driving high incidence and mortality rates. Amidst all this, LIRA’s TB detection model continues to demonstrate consistently high sensitivity across diverse and inclusive datasets, making it well-suited for deployment in community screening initiatives and primary health centers where specialist expertise is limited. The three-zone classification framework provides a pragmatic approach wherein: redzone cases indicate rapid referral for confirmatory molecular testing (CBNAAT/TrueNat), yellow-zone cases undergo further clinical assessment or repeat imaging, and green-zone cases are reassured. This risk-based triage strategy is designed to overcome the limitations of current TB screening methods. It was noted that while symptom-based screening lacks sensitivity for early or subclinical illness, universal chest radiography without triage remains resource-intensive. LIRA addresses this gap by identifying and prioritizing radiographs most likely to show active disease, enabling targeted molecular testing. In doing so, it reduces workload and turnaround time for radiologists and clinicians, especially in emergency and resource-limited settings. This significantly enhances the efficiency and cost-effectiveness of existing screening workflows.^20, 21^ Scalable deployment of LIRA may prove beneficial for achieving the stage of TB elimination across India’s diverse and heterogeneous healthcare system. Its minimal infrastructure requirements, being basic computing hardware and good internet connectivity, further align with India’s expanding mobile health and telemedicine initiatives. This ensures smooth deployment of the model in remote areas and supports easy scalability across the country. Moreover, integration with national TB information systems and databases could facilitate real-time surveillance, identification of high-risk groups, and continuous monitoring of screening outcomes.^22^ Additionally, LIRA supports contact tracing and screening in crowded settings such as households, schools, prisons, army camps, and medical facilities. Rapid AI-assisted screening of these high-risk groups can help detect secondary cases earlier and interrupt transmission, while the three-zone design guarantees the efficient use of confirmatory testing resources.^23^

### Risk Stratification and Rapid Referral Pathways

LIRA demonstrates high sensitivity for acute and potentially life-threatening conditions, which enables effective risk stratification and rapid referral. Its strong performance in detecting pleural effusion (99.1% sensitivity on NIH data) and pneumothorax (94.8% sensitivity) allows it to serve as an early warning system, prompting alerts for immediate therapeutic intervention. In core medical emergencies such as massive pleural effusion or tension pneumothorax, rapid identification is critical, and AI-assisted triage makes sure that critical findings like these are not delayed by routine reporting protocols.^8^ With respect to cardiomegaly, early detection leads to timely referral to cardiology and initiation of heart failure management protocols. With cardiovascular disease prevalence on the rise in India due to aging, hypertension, diabetes, and rheumatic heart disease, early intervention can significantly lower hospitalizations and mortality rates.^24^ LIRA’s capacity to detect cardiomegaly supports screening in high-risk groups and facilitates longitudinal monitoring. The horizontal deployment of a single AI system capable of detecting multiple pathologies offers clear operational advantages over disease-specific tools. This optimizes the radiology worklist and reduces turnaround times for several clinical scenarios at once, rather than focusing on a single disease. Therefore, implementing one integrated solution simplifies training, workflow integration, and quality assurance while providing broad screening coverage across a multiplicity of clinical scenarios.^8^

### Generalizability and Real-World Considerations

As evidenced by its consistent performance across geographically diverse clinical datasets (NIH, Montgomery, CheXpert, Shenzhen, SIIM-ACR, Jaypee), LIRA demonstrates generalizability across diverse populations, imaging equipment, and treatment settings. This breadth of validation addresses critical ethical concerns regarding dataset diversity and algorithmic bias. These discrepancies likely represent intrinsic differences between carefully curated research datasets and real-world clinical practices. Research datasets usually comprise high-quality images obtained under standardized protocols and interpreted by expert radiologists, whereas real-world data includes portable radiographs, variable positioning, motion artifacts, and reports generated under time constraints.^25^ The comparatively lower specificity of AI reporting may also be shaped by multiple contextual factors, such as higher disease prevalence, local reporting thresholds, widespread use of portable imaging, and population-specific characteristics such as prior tuberculosis, chronic lung disease, and malnutrition, resulting in radiographic variations. Despite this, LIRA maintained high sensitivity on Indian datasets, which aligns with its intended primary use as a screening and triage tool. In public health screening, prioritizing sensitivity is crucial. False negatives carry greater consequences than false positives, as missed cases may go unchecked and contribute to disease progression and transmission. Whereas false positives can be resolved through subsequent confirmatory testing.^17^ Further optimization of real-world performance may be achieved through ongoing refinement using more diverse Indian datasets, especially from district hospitals and primary health centers.

### Integration into Clinical Workflow and Human-AI Cooperation

Clinical impact cannot be guaranteed by technical validation alone. Successful implementation and adoption require seamless workflow integration, clinician acceptance, and clearly defined human-AI collaboration models.^26^ LIRA’s rapid and robust processing enables deployment at multiple points in the care delivery system, including primary health center screening, emergency department triage, outpatient pre-screening, and radiologist work list prioritization. Each application necessitates tailored workflows, training, and quality assurance protocols.^17^ Positioning AI as clinical decision support rather than autonomous diagnosis is crucial for ethical application and professional acceptance. Diagnostic autonomy and responsibility still remain with clinicians, while AI only contributes to speed, consistency, and prioritization.^27^ To further enhance clinician trust and usability, transparency must be maintained. This can be achieved through building more explainable AI models through objective methods like visual explanations, confidence scores, and feedback mechanisms.^28^

### Screening of Abnormalities and Triaging of Pathologies

In resource-limited settings like India, high-burden pulmonary and cardiac conditions such as tuberculosis, pneumonia, and interstitial lung diseases impose significant financial and logistical challenges on patients. The deployment of AI-based chest radiograph (CXR) triage systems at the primary care level offers a transformative approach to early screening and triaging protocols. This can facilitate rapid identification and prioritization of abnormalities in the general population during community outreach, thereby enabling efficient triaging of pathologies in emergency and hospital scenarios. These systems have the potential to guide the patients towards the most appropriate diagnostic and treatment pathways at the outset.^19, 29^ Such early interventions not only minimize unnecessary expenditures on advanced imaging or referrals but also reduce the burden of travel and alleviate the loss of income due to redundant hospital visits. This particularly helps the rural or low-income populations who may otherwise endure prolonged waits and visits to tertiary centers. LIRA’s high sensitivity and specificity could support scalable implementation to alleviate overall healthcare system pressures and improve equitable access to timely care.

### Limitations and Future Directions

The applicability of this study is limited by its retrospective design; prospective trials are necessary to evaluate real-world impact on turnaround times, referral patterns, treatment initiation, and patient outcomes. Strong proof of therapeutic benefit and cost-effectiveness would come from randomized studies comparing AI-augmented and traditional workflows.^25,30^ Future research should also focus more on implementation challenges, economic analyses, and clinician acceptance. Continuous model training and refinement using diverse Indian data and research into optimal human-AI collaboration will be critical for safe, effective, and scalable deployment.

## CONCLUSION

LIRA exhibits multi-label performance across various datasets with consistently high sensitivities. This data strongly supports our model’s intended role as an AI-based chest radiograph screening and triage tool. This clinical validation study highlights the potential of deep-learning-based AI models to address critical diagnostic gaps in India. LIRA can potentially reduce diagnostic delays, enable early detection of preventable diseases by facilitating rapid, reliable screening, risk stratification, and prioritization of high-risk cases, and encourage timely referrals to appropriate levels of care within the healthcare delivery system. For tuberculosis screening, its high sensitivity and three-zone risk-based framework make it suitable for community outreach and primary healthcare centers. This aspect directly aligns with the national TB elimination priorities and strategies while simultaneously optimizing resource utilization. However, retrospective validation alone is not sufficient for adequate adoption. Future prospective clinical studies are required to establish the real-world impact of LIRA on patient outcomes, workflow integration, and cost-effectiveness of this intervention. Successful implementation will depend on appropriate clinician training, workflow integration tailored to specific needs, steady monitoring of the performance delivered, and regulatory oversight. Moreover, to address the observed specificity variations in this study, continuous refinement of the model using diverse Indian datasets will be required. Lastly, with suitable governance and human-AI collaboration, systems such as LIRA have the potential to expand screening and diagnostic capacity, address the burden of healthcare disparities, and meaningfully contribute to India’s public health mission.

## ACKNOWLEDGEMENTS

The authors express sincere gratitude to the developers of the public and open-access datasets used in this study. We acknowledge the work of the National Institutes of Health (NIH) for the Chest X-ray dataset, the Stanford CheXpert team, the Society for Imaging Informatics in Medicine - American College of Radiology (SIIM-ACR) Pneumothorax dataset collaborators, Jaypee University of Information Technology, and the teams behind the Montgomery County and Shenzhen datasets from the U.S. National Library of Medicine. These openly available resources were instrumental in validating our AI model. The authors also acknowledge the contributions of the Lenek Technologies team, including AI developers, engineers, and data scientists who supported model development, deployment, and technical aspects of the validation process. The datasets used in this study are publicly available from the repositories cited.

## DECLARATIONS

### Ethical approval

Ethical review and approval were waived for this study due to the nature of the study design, which involved the retrospective analysis of exclusively publicly available, de-identified chest radiograph datasets (National Institutes of Health (NIH) for the ChestX-Ray8 dataset, the Stanford CheXpert dataset, the Society for Imaging Informatics in Medicine - American College of Radiology (SIIM-ACR) Pneumothorax dataset, Jaypee University of Information Technology dataset, and the Montgomery and Shenzhen datasets from the U.S. National Library of Medicine).

### Data Availability Statement

All data used in this study are publicly available. The de-identified chest radiograph datasets analyzed during the current study are available in the following public repositories; National Institutes of Health (NIH, USA) ChestX-ray8 dataset (Available at: https://nihcc.app.box.com/v/ChestXray-NIHCC/folder/36938765345); Stanford Hospital (USA) CheXpert dataset (Available at: https://aimi.stanford.edu/datasets/chexpert-chest-x-rays); U.S. National Library of Medicine Montgomery County and Shenzhen datasets (Available at: https://data.lhncbc.nlm.nih.gov/public/Tuberculosis-Chest-X-ray-Datasets/index.html); Society for Imaging Informatics in Medicine - American College of Radiology (SIIM-ACR) Pneumothorax dataset (Available at: https://www.kaggle.com/c/siim-acr-pneumothorax-segmentation); Jaypee University of Information Technology (India) Tuberculosis dataset (Available at: https://www.kaggle.com/datasets/raddar/chest-xrays-tuberculosis-from-india).

